# Epidemiological Philosophy of Pandemics

**DOI:** 10.1101/2021.02.24.21252304

**Authors:** Tareef Fadhil Raham

## Abstract

**Objectives:** Current estimates of the total number of cases of COVID-19 are largely based on previously-determined case fatality rates (CFRs). In this study, we aim to find an association between the Covid-19 number of cases / million inhabitants (M) and mortality rate (MR), and the association of Covid MR −19 and CFR. The background theory in this study is based on two factors: (1) There is no evidence that the CFR is fixed throughout time or place during an epidemic and (2) there is evidence that an increased viral load (density of infection) leads to more fatalities.

**Study Design:** We chose 31 countries with testing coverage levels of > 400,0000 tests /M and populations greater than 1 million inhabitants.

**Methods:** We used ANOVA regression analyses to test the associations.

**Results:** There was a very highly significant correlation between MR and the total number of cases/ million population inhabitants(M) (P-value 0.0000).

The CRF changed with a change in the MR. A very high positive influence of the COVID-19 MR on the CFR (P-value = 0.0002).

**Conclusions:** Increased number of cases per million inhabitants is associated with increased MR. Increased MR is associated with increased CFR. This finding might explain variable mortality rates that happened during this pandemic and possibly previous pandemics. This evidence will give us an idea of the behavior of epidemics in general.

## Introduction

There is strong evidence from various studies on the importance of the dose of the inoculum of a pathogen that can lead to severe infection.^1^ Such pathogens include those causing influenza ^2,3^ and the measles^4^, as well as Human Immunodeficiency Virus (HIV),^5^ tuberculosis (TB),^6, 7^ *Streptococcus pneumonia*,^*8*^ HBV^9^, flavivirus West Nile virus^10^, and Coronaviruses.^1,11^

The proposed mechanism by which a high viral inoculum leads to more severe disease is via a dysregulated and overwhelmed innate immune response to a higher viral dose, where immunopathology plays a role in viral pathogenesis ^12^. This may be the case for COVID-19.^13^

It has been suggested that a minimal viral inoculum may be controlled subclinically by innate defense mechanisms, while massive doses can overwhelm the innate immunity and may cause severe disease and rapid death. ^12^

Unfortunately, this issue has not undergone any challenge trials. Furthermore, epidemiological studies to correlate the association between the clustering of cases with both the mortality rate (MR) and the case fatality ratio (CFR) are lacking.

Several research groups have developed epidemiological models of COVID-19. These models use confirmed cases and deaths, testing rates, and a range of assumptions and epidemiological knowledge to estimate the number of true infections and other important metrics.^14^

CFR has gained great importance in the COVID-19 pandemic, because the expected total mortality burden of COVID-19 is directly related to the CFR. According to WHO, countries are making their final CFR estimates as active cases are resolved. Unfortunately, current CFR calculations during ongoing epidemics have been criticized due to the wide variation in CFR estimates over the course of an epidemic, making them difficult to compare for several reasons. These models might not accurately track the pandemic, as they apply previously determined infection fatality ratios (IFRs) from local sources or abroad. This makes currently used models using predetermined CFR subject to great bias. According to the World Health Organization (WHO), there has been broad variation in naïve estimations of CFR that may make them misleading. ^15,16,17^.

The testing capacity may be limited and restricted to people with severe cases of disease and priority risk groups. ^17^This makes continuous massive COVID-19 testing for continuous estimation of (IFR) or CRF a difficult task.

In the context of current variance and difficulties in CFR estimates and to shed light onto the unknown parameters associated with COVID-19 mortality, which are poorly understood, we investigated whether the CFR is associated with a change in MR expressed as the number of mortalities/million (M) inhabitant’s population. Moreover, this study also looks for an association and behavior of the MR expressed as deaths/M and the number of cases/M population, which (in high number settings) represents a high density of infection.

## Methods

Study design: This study was conducted to look for any relationship between the mortality rate (MR) presented as deaths/million (M) members of the population with both the total number of cases/(M) population (density of infection) and the CFR. We chose 31 countries with testing coverage of >400,0000 tests/M inhabitants and a population size of >1 million. Confirmed case counts adjustment were based on the percentage of COVID-19 tests that come back positive.

We used ANOVA regression analyses to test the associations measured throughout the study (SPSS-21)

Data were collected from the following public reference websites:

1. <u>COVID-19/Coronavirus Real Time Updates with Credible Sources in US and Canada”</u>. 1point3acres. Retrieved 16 January, 2021.
2. COVID-19 Dashboard by the Center for Systems Science and Engineering (CSSE) at Johns Hopkins University (JHU)”. ArcGIS. Johns Hopkins University.
3. WHO Coronavirus Disease (COVID-19) Dashboard https://covid19.who.int/?gclid=CjwKCAiA1eKBBhBZEiwAX3gql_TD3cUixzIROXwHFdS3yNhCxcF79FAYNtFC8mgHVXewA13pOFhEQxoCV1IQAvD_BwE
4. <u>COVID-19 Dashboard by the Center for Systems Science and Engineering (CSSE) at Johns Hopkins University (JHU)”. ArcGIS. Johns Hopkins University</u>
5. <u>COVID-19 Virus Pandemic—Worldometer (worldometers.info)</u>
6. <u>COVID-19 pandemic by country and territory—Wikipedia</u>

Information was collected on the number of COVID-19 cases/M inhabitants and the number of COVID-19 deaths/M inhabitants as at January 16, 2021. Additional country-specific references are included within the supplementary appendices.

The CFR was calculated by dividing the number of COVID-19 deaths up to January 16, 2021 by the number of confirmed cases up to that time, and this was expressed as a percentage. The MR was calculated by dividing the number of COVID-19 deaths per 1 M inhabitants.

### Statistical Methods

An optimum highly fitted model was checked among the several assumed non-linear regression models that can be transformed to linear equations, such as (logarithmic, inverse, quadratic, cubic, compound, power, S-shape, growth, exponential, and logistic). A simple linear regression model was proposed. Predicted equations have been suggested for studying the impact of “total cases per 1 M tested on Covid-19 death/1M population inhabitants concerning estimates (tables 1 and 2), and impact of “total Covid-19 death/1M population inhabitant on Covid-19 case fatality rate (table 3). All statistical methods were performed using the ready-made statistical package SPSS, ver. 21.

**Table (1):** Predicated equations for choosing an optimum highly fitted model.

| Predicated Equations | Models |
| --- | --- |
| Model whose equation is $Y = b_0 + (b_1 * t)$. | Linear |
| Model whose equation is $Y = b_0 + (b_1 * \ln(t))$. | Logarithmic |
| Model whose equation is $Y = b_0 + (b_1 / t)$. | Inverse |
| Model whose equation is $Y = b_0 + (b_1 * t) + (b_2 * t^{**2})$. | Quadratic |
| Model defined by the equation $Y = b_0 + (b_1 * t) + (b_2 * t^{**2}) + (b_3 * t^{**3})$. | Cubic |
| Model whose equation is $Y = b_0 * (t^{**b_1})$ or $\ln(Y) = \ln(b_0) + (b_1 * \ln(t))$. | Compound |
| Model whose equation is $Y = e^{**}(b_0 + (b_1/t))$ or $\ln(Y) = b_0 + (b_1/t)$. | S-Shape |
| Model whose equation is $Y = b_0 * (b_1^{**t})$ or $\ln(Y) = \ln(b_0) + (\ln(b_1) * t)$. | Power± |
| Model whose equation is $Y = 1 / (1/u + (b_0 * (b_1^{**t})))$ or $\ln(1/y-1/u) = \ln(b_0) + (\ln(b_1)*t)$ | Growth |
| Model whose equation is $Y = e^{**}(b_0 + (b_1 * t))$ or $\ln(Y) = b_0 + (b_1 * t)$. | Exponential |
| Model whose equation is $Y = b_0 * (e^{**}(b_1 * t))$ or $\ln(Y) = \ln(b_0) + (b_1 * t)$. | Logistic |

±Shaded predicated equations represents an optimal model.

**Table (2):** Impact of total cases to :1M. tested on total Covid-19 deaths / 1M.

| Dependent variable: "total deaths / 1million." |  |  |  |  |  |
| --- | --- | --- | --- | --- | --- |
| List wise deletion of missing data |  |  |  |  |  |
| Correlation Coefficient | 0.74800<br>(VHS) | Meaningful nonlinear regression<br>tested in a two tailed alternative<br>statistical hypothesis |  |  |  |
| R- Square | 0.5595 |  |  |  |  |
| F Statistic of Reg. ANOVA | 36.83451 | Sign. F = | 0.0000 (VHS) <sup>(*)</sup> |  |  |
| Variables in the Equation |  |  |  |  |  |
| Variable | B | SE.B | Beta | t-test | Sig. level |
| Total cases per 1 M tested | 1.010683 | 0.166528 | 0.747998 | 6.069 | 0.0000 (VHS) <sup>(*)</sup> |
| (Constant) | 0.010813 | 0.018544 | - | 0.583 | 0.5643(NS) <sup>(**)</sup> |
| Predicted equation - power-shape model |  |  |  |  |  |
| (Total deaths / 1M) = (0.010813) * (total cases / 1 M tested)<sup>1.010683</sup> |  |  |  |  |  |

<sup>(\*)</sup> VHS: Very highly Sig. at $P < 0.000$ ; testing non-linear regression (power model): model whose equation is $Y = b_0 * (x^{b_1})$ or $\ln(Y) = \ln(b_0) + [b_1 * \ln(x)]$ .
<sup>(\*\*)</sup> NS: $P > 0.05$

**Table (3):** Non- linear (power mode) regression for total Covid-19 deaths /1M inhabitants on case fatality rate.

| Dependent variable Method... power -shape Model "case fatality rate." |  |  |  |  |  |
| --- | --- | --- | --- | --- | --- |
| List wise deletion of missing data |  |  |  |  |  |
| Correlation Coefficient | 0.67256 (VHS) | Meaningful nonlinear regression tested in a two tailed alternative statistical hypothesis |  |  |  |
| R- Square | 0.37746 |  |  |  |  |
| F Statistic of Reg. ANOVA | 23.9527 | Sign. F = | 0.0000 (VHS) (***) |  |  |
| Variables in the Equation |  |  |  |  |  |
| Variable | B | SE.B | Beta | t-test | Sig. level<sup>(*)</sup> |
| Total deaths./1M. | 0.446412 | 0.091213 | 0.67256 | 4.894 | 0.0000 |
| (Constant) | 0.089347 | 0.048811 | - | 1.830 | 0.0775 |
| Predicted equation – power shape model |  |  |  |  |  |
| $(\widehat{\text{Case fatality rate}}) = (0.0932) * (\text{total deaths} / 1\text{M.})^{0.446412}$ | | | | | |

(\*\*\*) VHS: Very Highly Sig. at P<0.000; Testing Non Linear Regression (Power Model): Model whose equation is $Y = b_0 * (x^{b_1})$ or $\ln(Y) = \ln(b_0) + [b_1 * \ln(x)]$ .

## Results and Findings

Table (2) shows a meaningful nonlinear regression (power model) tested in a two-tailed alternative statistical hypothesis. The slope value indicated that with an increasing one unit of the total Covid-19 cases per 1 M tested persons, there is a positive impact on the unit of total Covid-19 deaths/1million population inhabitant, and estimated as (1.010683), which was recorded as a very highly significant at P-value<0.000, as well as the relationship coefficient which was accounted as (0.748) with a meaningful and significant determination coefficient (R-square = 55.95%). Another source of variations that was not included in the model (intercept) was not significant at P_-value_ >0.05. This indicates that the assignable factor (i.e. total Covid-19 cases per1 M tested persons) was explaining all variations among the studied function’s factor (i.e. the total deaths/1M population inhabitants).

Figure No. (1): Shows the long-term trend of scatter diagram impact of total Covid-19 cases per 1 M tested population inhabitants on total Covid-19 deaths / M population inhabitants.

**Figure (1):**
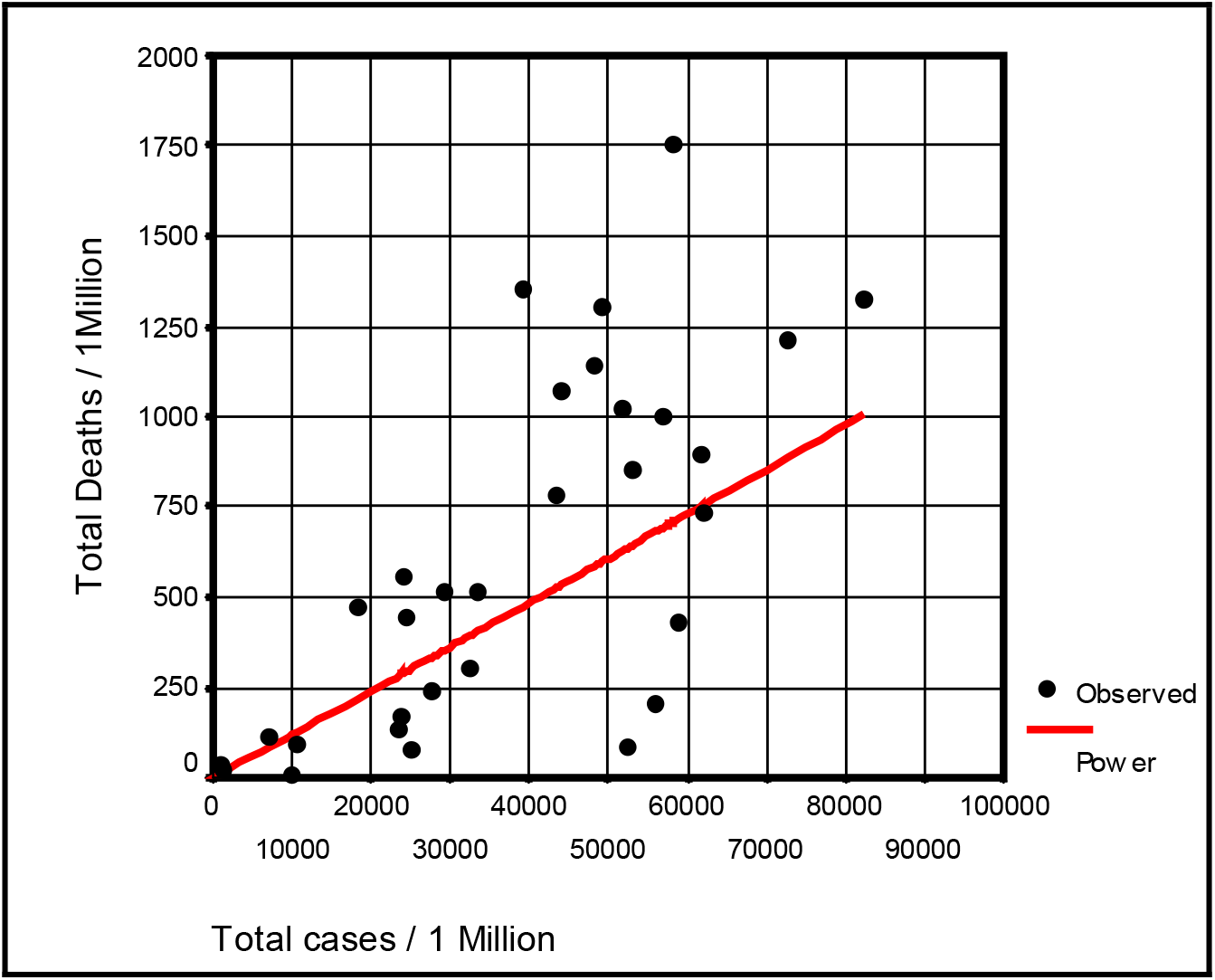
**Long term trend of the scatter diagram concerning impact of total Covid-19 cases per 1 M tested inhabitants on total Covid-19 deaths / 1M inhabitants.**

Table (3) shows a meaningful nonlinear regression (power model) tested in a two - tailed alternative statistical hypothesis. The slope value indicated that with an increasing one unit of the “total Covid-19 deaths /1M inhabitants “, there was a positive impact on the unit of “case fatality rate”, and was estimated as (0.446412), and recorded a very high significant impact at P-value<0.000. The relationship coefficient was accounted as (0.67256) with a meaningful and significant determination coefficient (R-Square = 45.234%). Another source of variations that was not included in the studied model, i.e. “intercept” showed no significant P_-value_ > 0.05. This indicates that the assignable factor (total Covid-19 deaths /1M inhabitants) explained all the variations among the studied function’s factor (i.e. the case fatality rate).

Figure No. (2): Shows the long term trend of scatter diagram impact of total Covid-19 deaths /1M inhabitants on case fatality rate.

**Figure (2):**
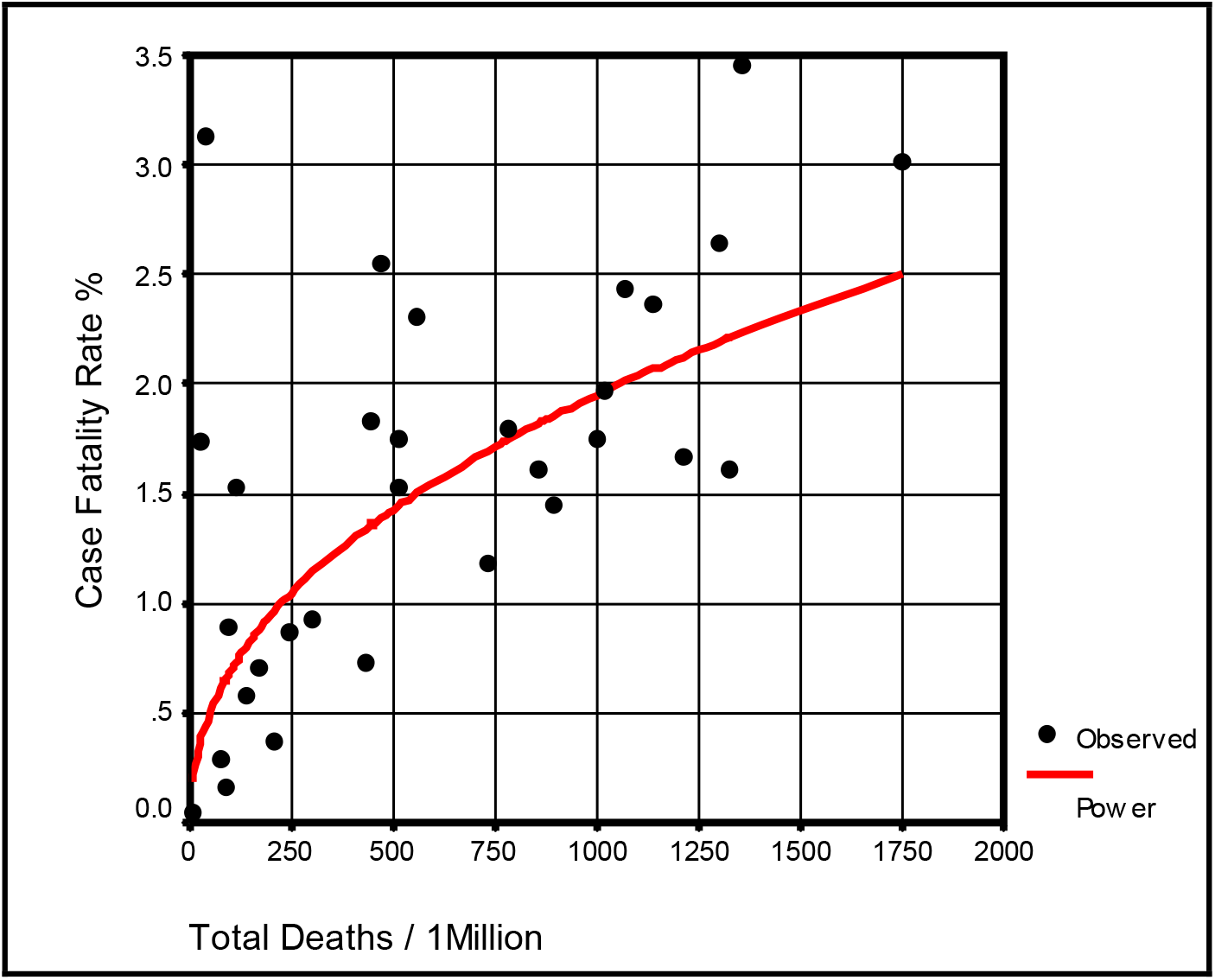
**Long term trend of the scatter diagram concerning impact of total Covid-19 deaths /1M inhabitants on case fatality rate**

## Discussion

In this study, we found that when an increase in number of Covid-19 cases /M inhabitants leads to increase inCovid-19 deaths/M in very high significant association (table 2 and figure1). We proved that when MR increases there is a concurrent increase in CFR in a very highly significant association too. According to these findings we suggest that a change in number of cases/M can modify current estimates of the CFR. By building models that are suitable for IFR we can predict the number of predicted mortalities according to total Covid-19 cases/M. We think that the suggested total number of cases as a factor determines MR and the suggested MR in determining CFR are novel findings in pandemics, behavior. Previous works that tried to find Covid-19 CFR did not take in consideration the number of cases among specified community. For example, the *Diamond Princess* cruise ship previous mortality estimates ^18,19,20^ possibly reflects behavior and CFR of Covid-19 concerning one condition, that is the total number of cases. So we think isolated estimates do reflect a specific behavior of Covid-19 pandemic concerning the specific number of cases studied in that community and not reflects the general behavior of pandemic, unless the number of cases is considered.

The results prove that the positive influence of the COVID-19 MR on the CFR and number of confirmed cases was very high by non-linear regression test (tables 3 and fig. 2). The most important confounder is the testing coverage and that the reported case counts were not adjusted to different levels of testing coverage and changing ratios of test positivity within time and place. Comparisons based on confirmed case counts can be misleading since a low case count may reflect either a low disease prevalence or a low rate of disease detection. Other confounding factors are unavoidable lags associated with most ways of measuring disease prevalence, and high prevalence of asymptomatic cases, which were estimated to be 10–70% of the total number of true infections elsewhere.^21^

On the other hand, estimates of the case fatality ratio (CFR) and infection fatality ratio (IFR) made in real time can be biased upwards by the under-reporting of cases and downwards by failure to account for the delay from confirmation-to-death, misattribution of deaths to other causes, and to limited access to testing.

Factors such as mobility, social distancing policies, population density, and host factors can interfere greatly with the number of cases. The scope of this study was not designed to look for causes concerning variance in the number of cases at different times and in different places. Instead, we test how the total number of cases affects the MR and how MR affects CFR. The number of cases, as a factor, determines the MR is well known, but the role of the number of cases in initiating cascade leading to increase CFR is a novel finding. The underlying hypothesis might be explained by the viral load and density of infection that are associated with increase in density and clusters of cases. Although this has been studied under certain conditions previously, the contribution of the number of cases in disease pathogeneses should be examined in depth again. During this pandemic, many studies have tried to determine why CFR widely differs rather than looking at why the number of cases/M inhabitants differs. It was suggested, for example, that overdrive of innate immunity possibly initiated when the viral load is high. This could manifests clinically as severe COVID-19 disease or multisystem inflammatory syndrome (MIS-C), a Kawasaki disease (KD)-like syndrome that occur mainly in children^22,23,24,25^,^26^. Family and community clusters of severe COVID-19 infection have been reported early in this pandemic. ^27,28^

Reported excess deaths estimates were thought to represent misclassified COVID-19 deaths or potentially those indirectly related to the COVID-19 pandemic. It was suggested that this excess number was related to the pandemic itself and not to disease, i.e., it was attributed to lack of facilities during the pandemic.^29^ According to our estimates, this excess could represent a portion of the total deaths attributed to increase deaths/M inhabitants associated with increase in number of cases specially during sharp waves of pandemic.

Previously, it has not been easy to explain the extraordinarily high mortality rates during certain epidemics, and this has been a concern for scientists. For example, there have been high mortality rates during measles epidemics in the Pacific Islands, such as in Fiji in 1875 and Rotuma in 1911, with mortality rates of 20% and 13% of the total residents, respectively. The mortality rate in the Faroe Islands in 1846 was nearly 10 times higher than that during the 1911 epidemic in Rotuma.^30^ One of the amazing things in these epidemics is that there was no direct evidence of hypervirulent strains of the measles virus or genetic predispositions to fatal outcomes after measles infection. ^31^

This makes us consider the role of high number of cases in initiation of high MR which leads to high CFR.

Again, in the 1918 influenza epidemic, virulence was notable when the number of deaths exceeded 20 million worldwide, with approximately half a million of these occurring in the United States.^32^ Noteworthy evidence from the 1918 epidemic was that one-quarter of the American population had clinically recognizable cases of flu during the epidemic, giving the impression of a high attack rate.^33^ Before the 1918 epidemic, one has to go back to the black death (bubonic plague) of 1346 to find a similarly devastating epidemic.^32^

During the COVID-19 pandemic, clinicians have struggled to understand why some infected patients experience only mild symptoms while others exhibit progressive, fatal disease.^27^ I think this new evidence enable us to understand a great part of this amazing situation.

## Advantages and conclusions

This is for the first time MR is addressed to predict CFR. Furthermore, it is a first time the attack rate (cases/M) is addressed in pathogeneses of sever Covid-19 expressed as high MR leads to a high CFR.

These findings might explain previous pandemics and as well to current pandemic or future ones.

An increase in the number of deaths/M inhabitants coincides with increases in the number of cases/M inhabitants and the CFR. This means that, when such situation happens a proportional increase in the total number of deaths outnumbers the proportional increase in total infections, which leads to an increased CFR from its baseline level. The proxy indicator for the MR and CFT is an increase in the total number of cases.

When CFR starts to increase during epidemic, this means an accelerated sever phase is taking place.

These findings will help in the development of infection control policies to break the chain of the pandemic and help to understand the philosophy of the pandemic.

Health systems should focus on decreasing the number of total cases, since MR increased with an increase in the total number of cases, and CFR can increase when MR is high enough to cause proportionally higher mortalities in relation to affected cases.

We suggest that all health systems can have the high MR and CFR, if the number of cases/M increases.

Better understanding of the pandemic behavior through showing that an increased CFR with an increased number of cases supports the viral overload theory.

## Data Availability

data is available through supplementary attached file

## Conflict of interest

There are no conflicts of interest worth mentioning. There is no funding source.

## Acknowledgment

I am deeply grateful to Emeritus Professor Abdulkhaleq Abduljabbar Ali Ghalib Al-Naqeeb, Ph.D.in the Philosophy of Statistical Sciences speciality at the Medical & Health Technology college, Baghdad-Iraq, for his assistance and support with the data analysis, interpretation of the findings, and statistical revision of the paper.

Ethical approval was not required for this study, as we used publically available data, and patients were not involved.

## Notes

### Competing Interest Statement

The authors have declared no competing interest.

### Author Declarations

I used publically available data.Patients were not involved

### Summary of Updates

Two tables were revised using the same original data. The methodology was revised to clarify results, findings, and conclusions.

