## Supplementary file for "Epidemiological Philosophy of Pandemics"

Appendix 1 references for morbidity and mortality data

1- [**COVID-19/Coronavirus Real Time Updates With Credible Sources in US and Canada"**](https://coronavirus.1point3acres.com/en)**. 1point3acres.**

**Retrieved 16 January, 2021.**

2- [**COVID-19 Dashboard by the Center for Systems Science and Engineering (CSSE) at Johns Hopkins University (JHU)"**](https://gisanddata.maps.arcgis.com/apps/opsdashboard/index.html#/bda7594740fd40299423467b48e9ecf6)**.**

[**ArcGIS**](https://en.m.wikipedia.org/wiki/ArcGIS)**.** [**Johns Hopkins University**](https://en.m.wikipedia.org/wiki/Johns_Hopkins_University)**.**

3- [**WHO Coronavirus Disease (COVID-19) Dashboard**](https://covid19.who.int/)

[**https://covid19.who.int/?gclid=CjwKCAiA1eKBBhBZEiwAX3gql_TD3cUixzIROXwHFdS3yNhCxcF79FAYNtFC8mgHVXewA13pOFhEQxoCV1IQAvD_BwE**](https://covid19.who.int/?gclid=CjwKCAiA1eKBBhBZEiwAX3gql_TD3cUixzIROXwHFdS3yNhCxcF79FAYNtFC8mgHVXewA13pOFhEQxoCV1IQAvD_BwE)

Appendix 2

| **No.** | **Country** | **References for data collection** |
| --- | --- | --- |
| **1** | **USA** | [**COVID-19/Coronavirus Real Time Updates With Credible Sources in US and Canada"**](https://coronavirus.1point3acres.com/en)**.** |
| **2** | [**Russia**](https://srv1.worldometers.info/coronavirus/country/russia/) | **Reference (in Russian). Contact author for more information** |
| **3** | [**UK**](https://srv1.worldometers.info/coronavirus/country/uk/) | [**Coronavirus (COVID-19) in the UK"**](https://coronavirus.data.gov.uk/)**. coronavirus.data.gov.uk.** |
| **4** | [**France**](https://srv1.worldometers.info/coronavirus/country/france/) | **1- References (in French). Contact author for more information.**  [**www.santepublique**](http://www.santepublique) |
| **5** | [**Italy**](https://srv1.worldometers.info/coronavirus/country/italy/) | **[COVID-19 ITALY]. Reference (in Italian). Contact author for more information.** |
| **6** | [**Spain**](https://srv1.worldometers.info/coronavirus/country/spain/) | **References (in Spanish). Contact author for more information.** |
| **7** | [**Germany**](https://srv1.worldometers.info/coronavirus/country/germany/) | **References (in German). Contact author for more information.** |
| **8** | [**Czechia**](https://srv1.worldometers.info/coronavirus/country/czech-republic/) | **Contact author for more information.** |
| **9** | [**Canada**](https://srv1.worldometers.info/coronavirus/country/canada/) | [**Timeline of the COVID-19 pandemic in Canada - Wikipedia**](https://en.wikipedia.org/wiki/Timeline_of_the_COVID-19_pandemic_in_Canada#January_2)  **COVID-19 pandemic in Canada - Wikipedia** |
| **10** | [**Belgium**](https://srv1.worldometers.info/coronavirus/country/belgium/) | **Reference in (in Dutch). Contact author for more information.** |
| **11** | [**Israel**](https://srv1.worldometers.info/coronavirus/country/israel/) | **[Corona virus in Israel] (in Hebrew). Contact author for more information.** |
| **12** | [**Portugal**](https://srv1.worldometers.info/coronavirus/country/portugal/) | **Reference (in Portuguese). Contact author for more information.** |
| **13** | [**Sweden**](https://srv1.worldometers.info/coronavirus/country/sweden/) | **Reference (in Swedish). Contact author for more information.** |
| **14** | [**Switzerland**](https://srv1.worldometers.info/coronavirus/country/switzerland/) | [**Current situation in Switzerland"**](https://www.bag.admin.ch/bag/en/home/krankheiten/ausbrueche-epidemien-pandemien/aktuelle-ausbrueche-epidemien/novel-cov/situation-schweiz-und-international.html)**. Federal Office of Public Health.** |
| **15** | [**Austria**](https://srv1.worldometers.info/coronavirus/country/austria/) | [**Refference**](https://www.ots.at/pressemappe/54/bundesministerium-fuer-inneres) **(in German). Contact author for more information.** |
| **16** | [**UAE**](https://srv1.worldometers.info/coronavirus/country/united-arab-emirates/) | [**UAE CORONAVIRUS (COVID-19) UPDATES"**](https://covid19.ncema.gov.ae/en)**. National Emergency Crisis and Disaster Management Authority (UAE).** |
| **17** | [**Georgia**](https://srv1.worldometers.info/coronavirus/country/georgia/) | [**StopCOV.ge"**](https://stopcov.ge/en)**.** |
| **18** | [**Belarus**](https://srv1.worldometers.info/coronavirus/country/belarus/) | **Reference (in Russian). Contact author for more information.** |
| **19** | [**Denmark**](https://srv1.worldometers.info/coronavirus/country/denmark/) | **Reference (in Danish). Contact author for more information.** |
| **20** | [**Lithuania**](https://srv1.worldometers.info/coronavirus/country/lithuania/) | **Reference (in Lithuanian). Contact author for more information.** |
| **21** | [**Ireland**](https://srv1.worldometers.info/coronavirus/country/ireland/) | [**Latest updates on COVID-19 (Coronavirus)"**](https://www.gov.ie/en/news/7e0924-latest-updates-on-covid-19-coronavirus/)**.** [**Department of Health (Ireland)**](https://en.m.wikipedia.org/wiki/Department_of_Health_(Ireland))**.** [**Ireland's COVID-19 Data Hub - ICU, Acute Hospital & Testing Data"**](https://covid19ireland-geohive.hub.arcgis.com/pages/hospitals-icu--testing)**. gov.ie.** [**Department of Health**](https://en.m.wikipedia.org/wiki/Department_of_Health_(Ireland)) |
| **22** | [**Qatar**](https://srv1.worldometers.info/coronavirus/country/qatar/) | [**Coronavirus Disease 2019 (COVID-19)"**](https://covid19.moph.gov.qa/EN/Pages/default.aspx)**. Ministry of Public Health (Qatar).** |
| **23** | [**Bahrain**](https://srv1.worldometers.info/coronavirus/country/bahrain/) | [**Daily COVID-19 Report"**](https://healthalert.gov.bh/en/category/daily-covid-19-report)**. Ministry of Health (Bahrain)** |
| **24** | [**Singapore**](https://srv1.worldometers.info/coronavirus/country/singapore/) | [**COVID-19 Situation Report [Summary of Confirmed Cases by Status in the Past 14 Days]"**](https://covidsitrep.moh.gov.sg/)**.** [**Ministry of Health (Singapore)**](https://en.m.wikipedia.org/wiki/Ministry_of_Health_(Singapore))**.** |
| **25** | [**Norway**](https://srv1.worldometers.info/coronavirus/country/norway/) | **Contact author for more information.** |
| **26** | [**Latvia**](https://srv1.worldometers.info/coronavirus/country/latvia/) | **[Distribution of Covid-19 in Latvia].  (in Latvian).**  **Contact author for more information.** |
| **27** | [**Finland**](https://srv1.worldometers.info/coronavirus/country/finland/) | [**Situation update on coronavirus [Finland's situation in brief]"**](https://thl.fi/en/web/infectious-diseases-and-vaccinations/what-s-new/coronavirus-covid-19-latest-updates/situation-update-on-coronavirus)**. Finnish Institute for Health and Welfare. 22 February 2021**  **Contact author for more information.** |
| **28** | [**Estonia**](https://srv1.worldometers.info/coronavirus/country/estonia/) | [**Information about Coronavirus disease COVID-19"**](https://koroonakaart.ee/en)**. Estonian Health Board.**  **Contact author for more information.** |
| **29** | [**Australia**](https://srv1.worldometers.info/coronavirus/country/australia/) | [**Coronavirus (COVID-19) current situation and case numbers"**](https://www.health.gov.au/news/health-alerts/novel-coronavirus-2019-ncov-health-alert/coronavirus-covid-19-current-situation-and-case-numbers)**. Australian Government Department of Health.** |
| **30** | [**Cyprus**](https://srv1.worldometers.info/coronavirus/country/cyprus/) | **1-**[**COVID-19 Spread in Cyprus"**](https://covid19.ucy.ac.cy/)**. covid19.ucy.ac.cy. 21 February 2021.** |
| **31** | [**Hong Kong**](https://srv1.worldometers.info/coronavirus/country/china-hong-kong-sar/) | [**Coronavirus Disease (COVID-19) in HK"**](https://chp-dashboard.geodata.gov.hk/covid-19/en.html)**. Hong Kong: Department of Health.** |

Appendix 3 data collection with correction pf total cases to 1 million tested population

| **indicator** | **Total cases /1 M** | **Total deaths/1M** | **Case fatality rate %** | **Tests /1M** |
| --- | --- | --- | --- | --- |
| **Country** |  |  |  |  |
| **USA** | **72,654** | **1,211** | **1.66** | **849,381** |
| [**Russia**](https://srv1.worldometers.info/coronavirus/country/russia/) | **24,283** | **446** | **1.83** | **659,046** |
| [**UK**](https://srv1.worldometers.info/coronavirus/country/uk/) | **49,315** | **1,301** | **2.638** | **927,432** |
| [**France**](https://srv1.worldometers.info/coronavirus/country/france/) | **43,961** | **1,070** | **2.433** | **600,343** |
| [**Italy**](https://srv1.worldometers.info/coronavirus/country/italy/) | **39,209** | **1,354** | **3.453** | **479,942** |
| [**Spain**](https://srv1.worldometers.info/coronavirus/country/spain/) | **48,160** | **1,140** | **2.367** | **615,318** |
| [**Germany**](https://srv1.worldometers.info/coronavirus/country/germany/) | **24,187** | **557** | **2.302** | **433,140** |
| [**Czechia**](https://srv1.worldometers.info/coronavirus/country/czech-republic/) | **82,456** | **1,326** | **1.608** | **507,378** |
| [**Canada**](https://srv1.worldometers.info/coronavirus/country/canada/) | **18,427** | **469** | **2.545** | **432,394** |
| [**Belgium**](https://srv1.worldometers.info/coronavirus/country/belgium/) | **58,112** | **1,752** | **3.014** | **648,070** |
| [**Israel**](https://srv1.worldometers.info/coronavirus/country/israel/) | **58,914** | **429** | **0.728** | **1,024,098** |
| [**Portugal**](https://srv1.worldometers.info/coronavirus/country/portugal/) | **52,985** | **855** | **1.613** | **613,699** |
| [**Sweden**](https://srv1.worldometers.info/coronavirus/country/sweden/) | **51,659** | **1,019** | **1.972** | **465,556** |
| [**Switzerland**](https://srv1.worldometers.info/coronavirus/country/switzerland/) | **56,995** | **998** | **1.751** | **457,903** |
| [**Austria**](https://srv1.worldometers.info/coronavirus/country/austria/) | **43,447** | **781** | **1.797** | **445,755** |
| [**UAE**](https://srv1.worldometers.info/coronavirus/country/united-arab-emirates/) | **25,094** | **74** | **0.294** | **2,334,430** |
| [**Georgia**](https://srv1.worldometers.info/coronavirus/country/georgia/) | **61,989** | **732** | **1.180** | **510,059** |
| [**Belarus**](https://srv1.worldometers.info/coronavirus/country/belarus/) | **23,661** | **166** | **0.701** | **448,670** |
| [**Denmark**](https://srv1.worldometers.info/coronavirus/country/denmark/) | **32,430** | **301** | **0.928** | **2,036,378** |
| [**Lithuania**](https://srv1.worldometers.info/coronavirus/country/lithuania/) | **61,701** | **894** | **1.448** | **666,170** |
| [**Ireland**](https://srv1.worldometers.info/coronavirus/country/ireland/) | **33,527** | **511** | **1.524** | **552,627** |
| [**Qatar**](https://srv1.worldometers.info/coronavirus/country/qatar/) | **52,386** | **88** | **0.167** | **468,132** |
| [**Bahrain**](https://srv1.worldometers.info/coronavirus/country/bahrain/) | **55,984** | **206** | **0.367** | **1,460,392** |
| [**Singapore**](https://srv1.worldometers.info/coronavirus/country/singapore/) | **10,056** | **5** | **0.049** | **993,517** |
| [**Norway**](https://srv1.worldometers.info/coronavirus/country/norway/) | **10,712** | **95** | **0.886** | **573,565** |
| [**Latvia**](https://srv1.worldometers.info/coronavirus/country/latvia/) | **29,389** | **513** | **1.745** | **549,766** |
| [**Finland**](https://srv1.worldometers.info/coronavirus/country/finland/) | **7,231** | **111** | **1.535** | **475,983** |
| [**Estonia**](https://srv1.worldometers.info/coronavirus/country/estonia/) | **27,649** | **241** | **0.871** | **531,858** |
| [**Australia**](https://srv1.worldometers.info/coronavirus/country/australia/) | **1,118** | **35** | **3.130** | **482,119** |
| [**Cyprus**](https://srv1.worldometers.info/coronavirus/country/cyprus/) | **23,602** | **137** | **0.580** | **727,899** |
| [**Hong Kong**](https://srv1.worldometers.info/coronavirus/country/china-hong-kong-sar/) | **1,262** | **22** | **1.743** | **776,245** |
